# Epidemic control using stochastic and deterministic transmission models: performance comparison with and without parameter uncertainties

**DOI:** 10.1101/2022.11.12.22282246

**Authors:** Julien Flaig, Nicolas Houy

**Affiliations:** EPIMOD; GATE CNRS

**Keywords:** epidemic control, emerging disease, stochastic models, deterministic models, stochastic effects, value of information, disease X

## Abstract

**Background:** The spread of infectious diseases can be modeled using deterministic models assuming a continuous population or stochastic models assuming a discrete population. A stochastic model can be approximated by its deterministic counterpart under some conditions, however deterministic models are unable to captures effects pertaining to the discrete nature of populations, for instance epidemic extinction. We look into the choice of a model – deterministic or stochastic – from the perspective of decision making. We are interested in the influence of parameter uncertainties and of the quality of the estimates used to inform decisions.

**Method:** We consider an emerging disease in a closed population whose spread can be modeled by a stochastic SIR model or its deterministic version. Our objective is to minimize the cumulative number of symptomatic infected-days over the course of the epidemic by picking a vaccination policy out of three available options. We consider four decision making scenarios: based on the stochastic model or the deterministic model, and informed or under parameter uncertainty. We also consider different sample sizes covering parameter draws, stochastic model runs, or both depending on the scenario. We estimate the average performance of decision making in each scenario and for each sample size.

**Results:** The model used for decision making has an influence on the picked policies. The best achievable performance is obtained with the stochastic model, knowing parameter values, and for a large sample size. For small sample sizes, the deterministic model can outperform the stochastic model due to stochastic effects, both in the informed and the uninformed cases. Starting with the deterministic model under uncertainty, resolving uncertainties brings more benefit than switching to the stochastic model in our example.

**Conclusion:** This article illustrates the interplay between the choice of a type of model, parameter uncertainties, and sample sizes. It points to issues to be carefully considered when attempting to optimize a stochastic model.

## 1 Introduction

Infectious diseases spread in populations made up of discrete units, and events pertaining to this spread (whether individual encounters, recoveries, deaths, births or immigration) are essentially random. Stochastic transmission models – in which the population is assumed to be discrete – take these features into account. But stochastic models do not lend themselves easily to mathematical analysis. As for numerical approaches, they can be resource intensive as they may require many Monte Carlo simulations of epidemic trajectories. Early modeling efforts were then focused on deterministic transmission models in which the population is modeled as a continuum whose dynamics are represented by differential equations. A deterministic model approximates its corresponding stochastic model “for large populations”. Loosely speaking, as the population increases, the proportion of individuals, say, getting infected at a given time converges to the probability that any particular individual gets infected at that time by the law of large numbers.

Importantly, absolute numbers of individuals do not matter in a deterministic model: populations can be normalized or rescaled without changing the qualitative behavior of the system. This is of course a limitation in cases where absolute numbers of individuals do matter. The most obvious and widely studied of such situations are those in which extinction is possible, whether upon introduction of a new pathogen in a population, or in the endgame of an eradication attempt [1–3]. For instance, in a deterministic framework, the basic reproduction number *R*_0_ tells whether an emerging infectious disease will be able to spread in a population (*R*_0_ > 1) or not (*R*_0_ < 1) irrespective of the initial number of infectious individuals, which may assume any positive real value. In a stochastic model, by contrast, an emerging infectious disease may still become extinct before causing an outbreak if *R*_0_ > 1 with a probability depending on the (discrete) number of initially infectious individuals [4]. Thus, there is a rich literature using stochastic models to estimate the probability of an outbreak or of eradication in different scenarios [3, 5–7]. Beside the possibility of extinction, absolute numbers of individuals also matter where control means come in discrete units (think of hospital beds), although such scenarios have been less studied [8].

As for comparisons of deterministic and stochastic approaches, many previous studies compared the respective ability of deterministic and stochastic models to account for observed patterns for different parameter values and population sizes [2, 9–13]. These problems were chiefly put in terms of population biology – typically: under what conditions does a deterministic approximation give an accurate picture of a stochastic system? The objective of the present article is to shift perspective from population biology (or dynamics) to the perspective of decision making. Deterministic models are more convenient to use, but what is the cost of making decisions based on a deterministic model where a stochastic model would be more appropriate from a “biological” perspective? This issue is closely related to the problem of decision making under uncertainty. Indeed, the probability of stochastic effects such as extinction depends on the value of parameters that are most often uncertain. So, what is comparatively more costly: parameter uncertainties or the inappropriate use of a deterministic model? In addition, the quality of policy performance estimates – and hence of decision making – depends on the number of Monte Carlo samples used to produce these estimates. When using a stochastic model, Monte Carlo samples include samples of stochastic epidemic trajectories. In the case of decision making under parameter uncertainty (whether based on stochastic or a deterministic model), samples include epidemic trajectories for different random draws of parameter values. What, then, is the influence of sample size on the respective performance of stochastic and deterministic models, with or without parameter uncertainties?

We will answer these questions using two generic SIR transmission models: a stochastic model and its deterministic counterpart. In our scenario, a new pathogen is introduced in a susceptible population and the objective is to control its spread through single or double dose vaccination given a number of available vaccine doses. We consider four possible scenarios for decision making: using a stochastic model or its deterministic version, and deciding under parameter uncertainty or knowing parameter values. Notice that in these scenarios, the level of available information about the disease is a matter of context, while the use of a deterministic or stochastic model is a modeling choice (although it may depend on the context, e.g. on computational or organizational constraints). Also, we do not intend to describe the control of a specific disease or to make specific policy recommendations, but to give a qualitative and intuitive understanding of how the choice of a deterministic or stochastic model may influence decision making, and of the interplay between this choice and the level of available information about model parameters.

Yuan et al. [14] compared the use deterministic and stochastic models for decision making in a similar way as we do: they determined optimal vaccination policies based on a stochastic and a deterministic model, and assessed the performance of both policies when applied to the stochastic system. They showed for what parameter values (assumed to be known to the decision maker) the difference in performance was more or less significant. We look at the problem from a different angle by considering the perspective of a decision maker endowed with a certain level of information about model parameters, and deciding whether to use a deterministic or a stochastic model and with how many Monte Carlo samples. Another more technical difference is that Yuan et al. [14] numerically integrate the master equation of the stochastic model using the method proposed by Jenkinson and Goutsias [15] while we use a version of the Gillespie algorithm [16] to generate samples and estimate means.

We emphasize that the present article is not concerned with optimization methods or optimal control of stochastic models. For this we refer to [14, 17–20]. We restrict ourselves to a limited number of available policies (three), so that their respective performances can easily be estimated exhaustively. Solving the optimization problem then simply consists in picking the best policy based on these estimates. This does not make our results any less relevant: they apply to decision making in any situation where stochastic effects may be critical, whatever the optimization methods used to make decisions.

Finally, it is worth mentioning the distinction between demographic and environmental stochasticity [12]. We only consider demographic stochasticity, that is the randomness of events involving discrete individuals. Environmental stochasticity refers to the fact that parameter values may be noisy, even assuming a continuous population. The results presented in this article do not so much revolve around the noise involved by stochasticity, but rather around effects pertaining to the discrete nature of populations (in our case, the possibility of extinction). Randomness is necessary for our analyses to be relevant, however it is not sufficient: we also need a discrete population. See Grandits et al. [21] for an example of decision making based on a model assuming a continuous population with random variations.

The remainder of the article is organized as follow. Our method is presented in Section 2: the disease transmission models and parameter uncertainties (Sections 2.1 and 2.2 respectively), how we assume decisions to be made in the four considered scenarios (Section 2.3), and the validation method used to assess the performance of decision making in each case (Section 2.4). The results are presented in Section 3. Section 3.1 provides a general understanding of the behavior of our stochastic transmission model and of its deterministic counterpart. Section 3.2 shows the performance of decision making based on our stochastic model and its deterministic version under parameter uncertainty. Section 3.3 shows the same but assuming that parameter values are known to the decision maker. In Section 3.4, we compare decision making in all four scenarios: based either on the stochastic or deterministic model, and either under parameter uncertainty or knowing parameter values. Section 4 concludes.

## 2 Method

### 2.1 Disease transmission models and intervention policies

We consider a total population of *n* = 100 individuals, which corresponds to a hospital ward or a nursing home. The population is assumed to be closed. Individuals can be susceptible, infected and infectious, or recovered. Individuals may receive one, two, or no dose of vaccine. We assume that there is no waning of vaccination protection. We assume homogeneous mixing. All individuals get infected with contact rate *β* and they recover at rate *γ* – the basic reproduction number is *R*_0_ = *β/γ*. However, individuals who received *i* ∈ {1, 2} vaccine doses are less susceptible by a factor *σ*_*i*_ and less infectious by a factor *ρ*_*i*_ compared to unvaccinated individuals. Individuals who received *i* ∈ {0, 1, 2} doses of vaccine have have a probability *p*_*i*_ of developing symptoms. Model parameters are summarized in Table 1.

**Table 1:** Transmission model parameters.

| Parameter | Description |
| --- | --- |
| $R_0$ | Reproduction number of unvaccinated individuals. |
| $\gamma$ | Rate of recovery. |
| $\rho_1$ | Relative infectiousness of individuals having received 1 dose of vaccine. |
| $\rho_2$ | Relative infectiousness of individuals having received 2 doses of vaccine. |
| $\sigma_1$ | Relative susceptibility of individuals having received 1 dose of vaccine. |
| $\sigma_2$ | Relative susceptibility of individuals having received 2 doses of vaccine. |
| $p_0$ | Probability of symptomatic infection of unvaccinated individuals. |
| $p_1$ | Probability of symptomatic infection of individuals having received 1 dose of vaccine. |
| $p_2$ | Probability of symptomatic infection of individuals having received 2 doses of vaccine. |

At time 0, one person is infected and the others are susceptible. We assume that this initial case was detected, and our objective is to minimize the cumulative number of symptomatic infection-days over the course of the disease. At time 0, 50 vaccine doses are available and can be administered either as double doses to 25 individuals, or as single doses (to 50 individuals). We denote these policies “0-50” and “50-0” respectively. We also leave the possibility not to implement vaccination (policy “0-0”), even though this policy should be dominated. The set of available policy options is then *A* = {0-0, 0-50, 50-0}.

A policy is picked based either on a deterministic compartmental model *M*_0_, or on its stochastic counterpart *M*_1_. The compartmental models are described in more details in Appendix A. In the following, with a slight abuse of notation, we will use the same notations for deterministic and random variables.

### 2.2 Parameter uncertainties

We assume that the structure of the model is known as well as the number of initially infected individuals. Under parameter uncertainty, other parameters are uncertain but prior distributions are available to the decision maker. The parameter prior distributions are given in Appendix B and Figure 1 shows 512 draws from these distributions. For the sake of simplicity, we restrict *p*_0_ = 1. In the base case, we also fix *γ* = 0.1 so that all uncertainty regarding the spread of disease without intervention is summarized by uncertainty about a single parameter (*R*_0_ or *β*). All other uncertainties concern the respective levels of protection offered by single and double dose vaccination. Naturally, we impose that *p*_1_ *> p*_2_, *ρ*_1_ *> ρ*_2_ and *σ*_1_ *> σ*_2_.

**Figure 1:**
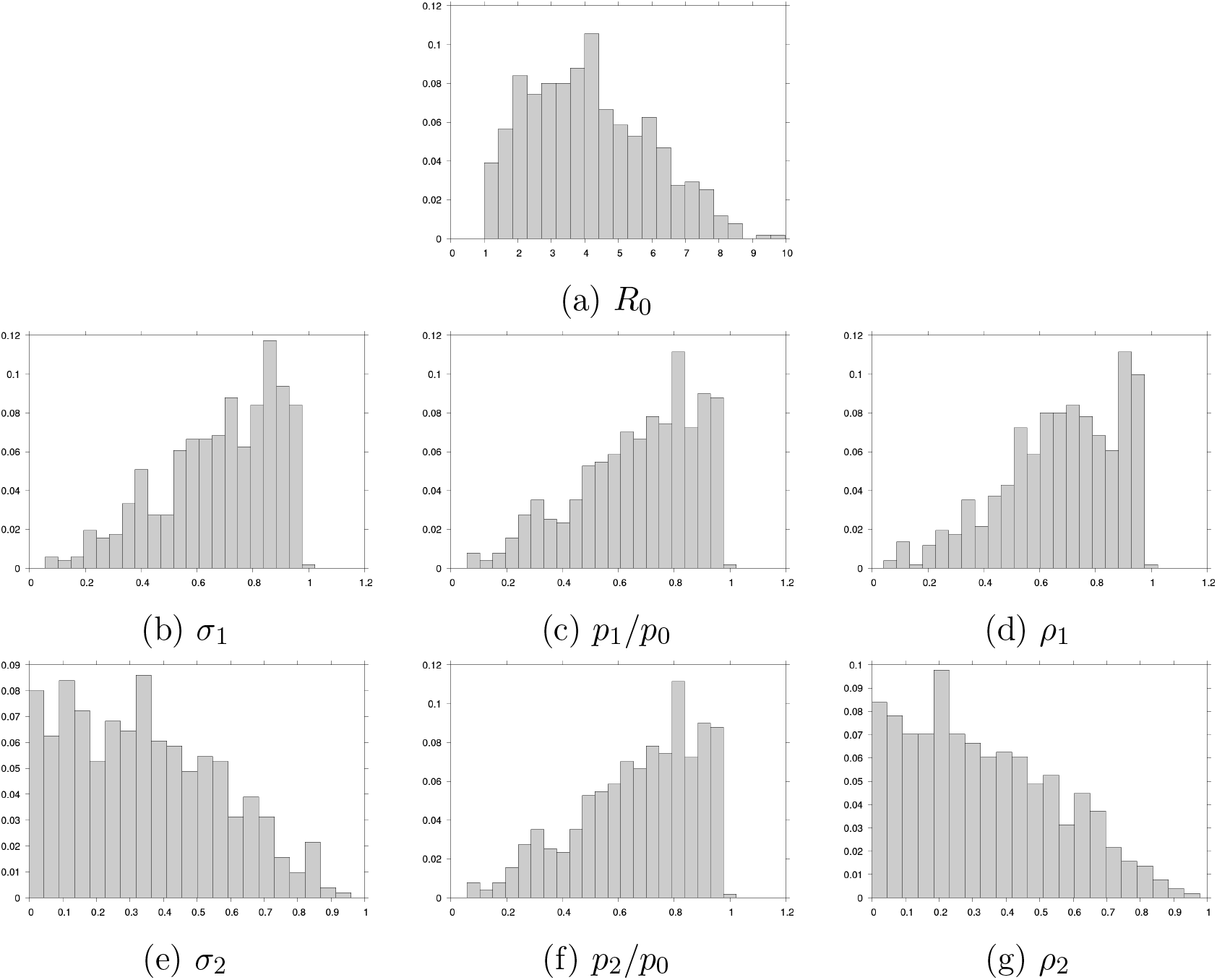
512 draws from the parameter prior distributions.

### 2.3 Decision making

Given parameter values *ξ*, policy *a* ∈ *A* can be evaluated based on deterministic model *M*_0_ or based on stochastic model *M*_1_. Let *C*_*i*_(*a, ξ*) the cumulative number of symptomatic infected-days over the course of the epidemic estimated with model *M*_*i*_. *C*_0_(*a, ξ*) is obtained directly by solving deterministic model *M*_0_. In the stochastic framework, *C*_1_(*a, ξ*) = *E* [*C*(*a, ξ*)], where *C* is a random variable – the number of symptomatic infected-days obtained with a single run of *M*_1_. *C*_1_ is estimated as the average of realizations of *C* over a number of runs of *M*_1_. This number will be varied in our analyses.

Under uncertainty about parameters *ξ*, policy 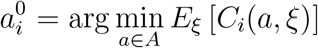 is then picked based on model *M*_*i*_: a policy is picked to minimize the expected number of symptomatic infected-days over the prior distribution of parameters. If the true value of *ξ* is known, policy 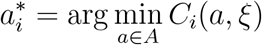 is picked based on model *M*_*i*_.

### 2.4 Validation of chosen policies

We assume that stochastic model *M*_1_ is an accurate representation of reality. Thus, given parameter values *ξ*, the outcome of chosen policy *a* once implemented is the result *C*(*a, ξ*) of a single stochastic epidemic trajectory of *M*_1_. We evaluate the performance *a priori* of policy *a* by estimating *E*_*ξ*_ [*C*_1_(*a, ξ*)] = *E*_*ξ*_ [*E* [*C*(*a, ξ*)]], the expected outcome over possible parameter values and epidemic trajectories. This performance can also be expressed in terms of averted symptomatic infected-days compared to the no vaccination policy: *E*_*ξ*_ [*C*_1_(0-0, *ξ*) *− C*_1_(*a, ξ*)]. Estimating the performance of decisions made under different scenarios – parameter uncertainties, type of model, Monte Carlo sample size – allows to compare the cost of uncertainties to the cost of using the “wrong” model or a too small sample size.

## 3 Results

### 3.1 Stochastic effects

Figure 2 illustrates the spread of the disease for each of the three available policy options, as estimated with deterministic model *M*_0_ and stochastic model *M*_1_, for each of the 512 parameter draws shown in Figure 1. For each parameter draw *ξ* (y-axis), a green dot shows the cumulative number *C*_0_(*a, ξ*) of symptomatic infected-days under policy *a* ∈ *A* estimated with deterministic model *M*_0_. Parameter draws are ordered by increasing *C*_0_(*a, ξ*). The heat map shows the distribution of the outcomes *C*(*a, ξ*) of 2,048 runs of stochastic model *M*_1_ for each parameter draw *ξ* and policy *a*. White dots show the average *C*_1_(*a, ξ*) of the 2,048 outcomes.

**Figure 2:**
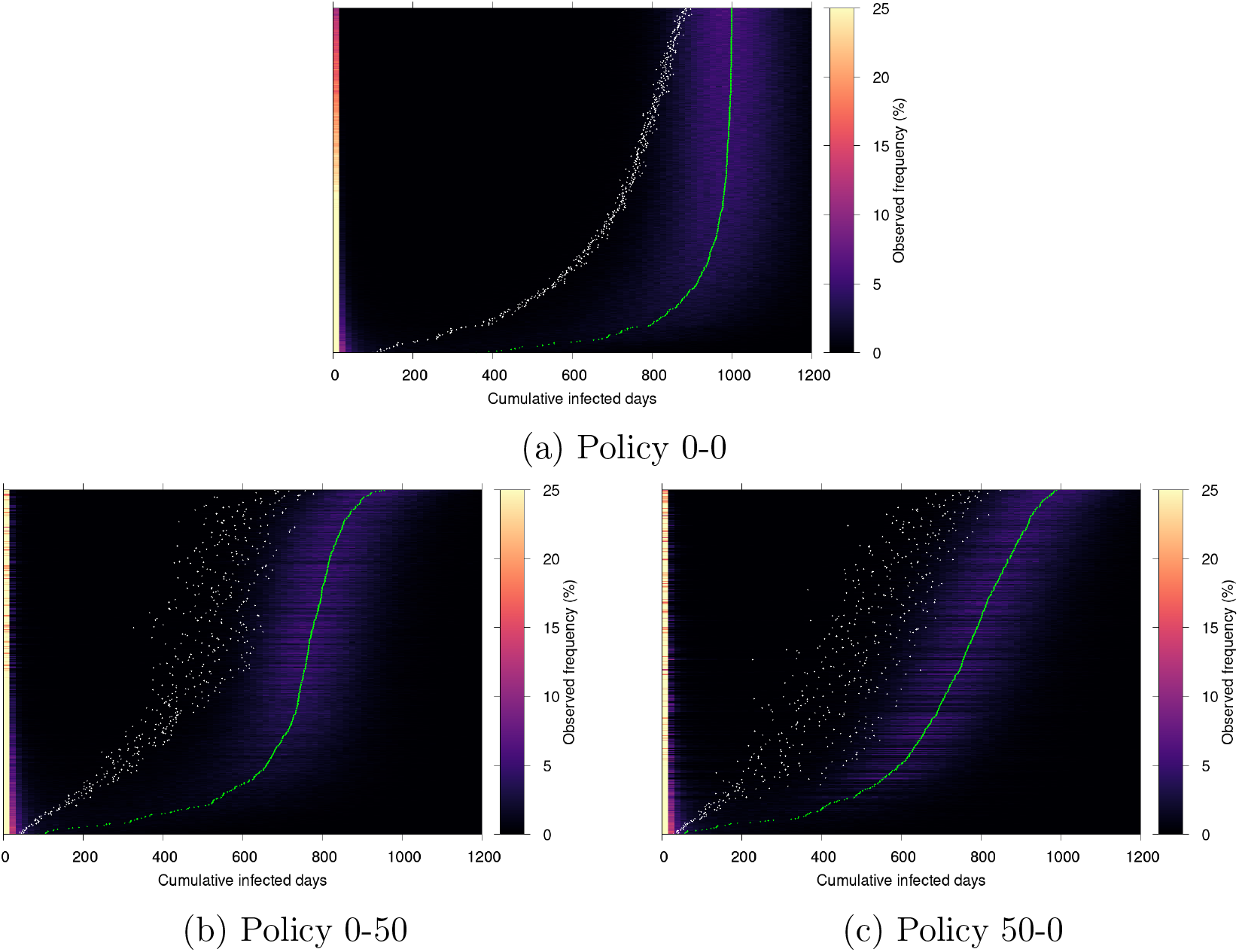
Observed frequency (over 2,048 stochastic simulations, color) of cumulative infected-days (x-axis) for each of the 512 data draws from the base case prior parameter distributions (y-axis). Green dots: outcomes of the deterministic simulations. White dots: averages of the stochastic simulations.

The distribution of *C*(*a, ξ*) is bimodal: in a portion of stochastic epidemic trajectories the disease goes extinct before spreading to a large part of the population (clear vertical bands on the left of the heat maps), while in others it causes a major outbreak (purple areas). This is a well-known feature of stochastic disease transmission models [3, 5, 7, 22]. Conditionally on the epidemic taking off, the deterministic model *M*_0_ gives a pretty good approximation (*C*_0_(*a, ξ*), green) of the of the average stochastic outcome (for each vaccination policy, the green dots are aligned with the purple areas of the heat maps). However the average stochastic outcome (*C*_1_(*a, ξ*), white) is dragged down by early extinction in some trajectories. By construction, these stochastic effects are not properly taken into account by the deterministic model *M*_0_.

### 3.2 Decision making under parameter uncertainty

Whether stochastic effects are overlooked or properly taken into account influences decision making. This is shown in Table 2 in the case of decision making under parameter uncertainty. Recall that under parameter uncertainty, policy 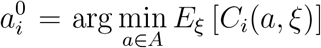 is picked based on model *M*_*i*_. If a decision is made based on deterministic model *M*_0_, policy is picked. If a decision is made based on stochastic model *M*_1_, policy 0-50 is picked. Once implemented, the former yields a total of about 453 symptomatic infected-days on average, and the latter about 439 symptomatic infected-days on average.^1^

**Table 2:** Estimate expected outcome of the three available policy option based on *M*_0_ and *M*_1_. Averages over 512×2,048 = 2^20^ simulations. In the stocastic case: 512 parameter draws, and 2,048 runs of *M*_1_ per parameter draw.

| Policy $a$ | $E_\xi [C_0(a, \xi)]$ estimate | $E_\xi [C_1(a, \xi)]$ estimate |
| --- | --- | --- |
| 0-0 | 941.371 (95% CI: 941.169 – 941.574) | 681.479 (95% CI: 665.501 – 697.457) |
| 0-50 | 726.308 (95% CI: 726.022 – 726.593) | 439.498 (95% CI: 424.672 – 454.324) |
| 50-0 | 726.012 (95% CI: 725.646 – 726.378) | 452.722 (95% CI: 435.771 – 469.673) |

This difference is small, but it still invalidates the naive reasoning according to which the deterministic model “assumes the worst” by ignoring the possibility of early epidemic extinction (in the stochastic sense: number of infectious individuals equal to exactly zero), hence leading to more conservative and thus effective policies. The edge of *M*_1_ over *M*_0_ is better understood from Figure 3, which shows the average (over parameter draws *ξ*) estimate cumulative density function of *C*(*a, ξ*). The two inflection points correspond to the two modes of the *C*(*a, ξ*) distributions shown in Figure 2. From the zoom in the right panel, we can see that policy 0-50 is better than policy 50-0 at avoiding the epidemic to take off. This can only be shown by modeling stochastic effects properly using stochastic model *M*_1_. Similarly, model *M*_1_ shows that policy 0-50 is better at avoiding the largest outbreaks: the proportion of outbreaks above 900 symptomatic infected-days is lower under policy 0-50 than under policy 50-0. Deterministic model *M*_0_, by contrast, considers a single possible trajectory per set *ξ* of parameter values, and predicts that on average over possible parameter values policies 0-50 and 50-0 will yield about 718 and 713 symptomatic infected-days respectively (Table 2). A vaccination policy may i) avoid the outbreak of the epidemic, or ii) limit the number of cases given an outbreak. Using a deterministic model for decision making means dealing with only the second effect.

**Figure 3:**
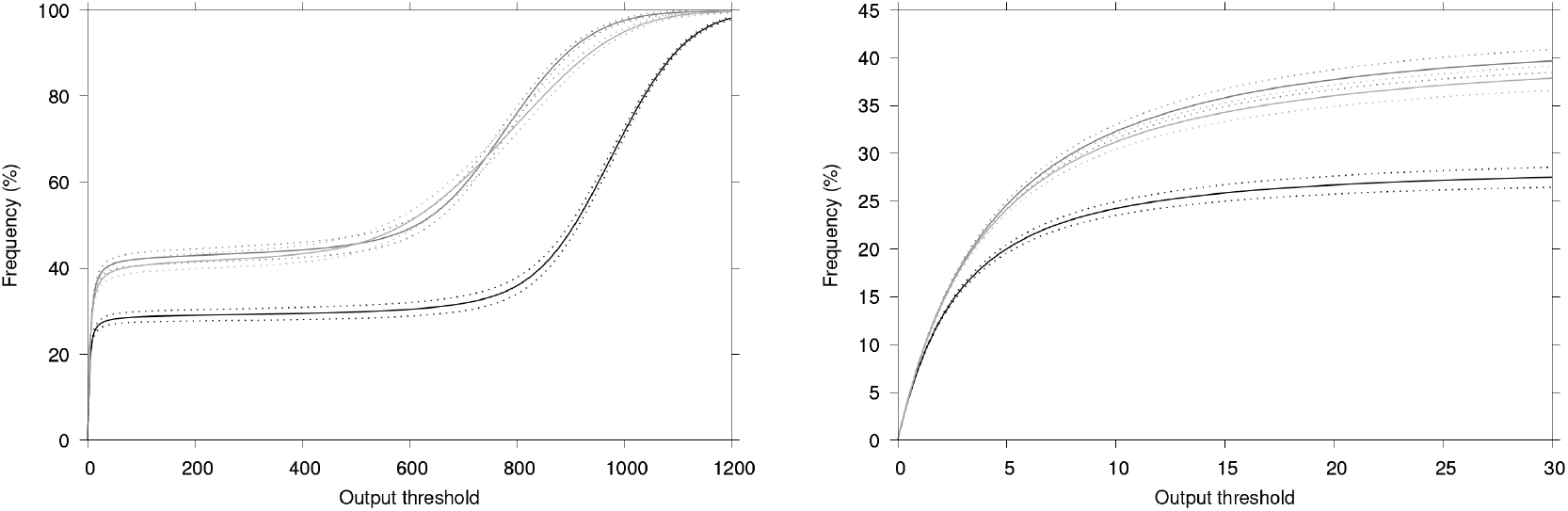
Average (over 512 data draws) frequency (over 2,048 stochastic simulations) of simulations yielding less cumulative symptomatic infected-days than the x-axis threshold value. Right: zoom for small values. Black: policy 0-0 (no vaccination), dark grey: policy 0-50, light grey: policy 50-0. Dotted lines: 95% CI.

So far, we assumed that policies were picked based on 512 parameter draws and, in the stochastic cases, 2,048 runs of stochastic model *M*_1_ for each parameter draw. Obviously, the number of parameter draws and of runs of *M*_1_ influences the quality of decision making. To illustrate this, we simulate decision making based on models *M*_0_ and *M*_1_ under parameter uncertainty and for varying numbers *N* of model simulations used to estimate *E*_*ξ*_ [*C*_0_(*a, ξ*)] and *E*_*ξ*_ [*C*_1_(*a, ξ*)] for each policy *a* ∈ *A*. When making a decision based on deterministic model *M*_0_, *N* is the number of parameter draws (the deterministic model is solved once for each draw). When making a decision based on stochastic model *M*_1_, *N* is the number of parameter draws and trajectory simulations – we bootstrap our 512 × 2,048 parameter draws and stochastic runs of *M*_1_. Each simulation of decision making is repeated 65,536 times for validation. We look at how often each policy is picked out of the 65,536 decision making simulations, and at the average performance of picked policies. Our bootstrapping procedures are detailed in Appendix D.

The results are shown in Figure 4. When a policy is chosen based on deterministic model *M*_0_ (Figure 4a), larger *N* means a better sampling of prior parameter distributions allowing for less volatile decision making. As *N* increases, decision 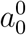 converges towards policy 50-0, which is 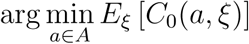 with a slight advantage over policy 0-50, as suggested by Table 2. By contrast, for *N* = 2, policies 0-50 and 50-0 are picked with almost equal probability, depending on the drawn parameter values. Since policies 0-50 and 50-0 have a close performance once implemented (see *E*_*ξ*_ [*C*_1_(*a, ξ*)] in Table 2), the average performance of decisions made based on *M*_0_ does not much depend on *N* (Figure 4b).

**Figure 4:**
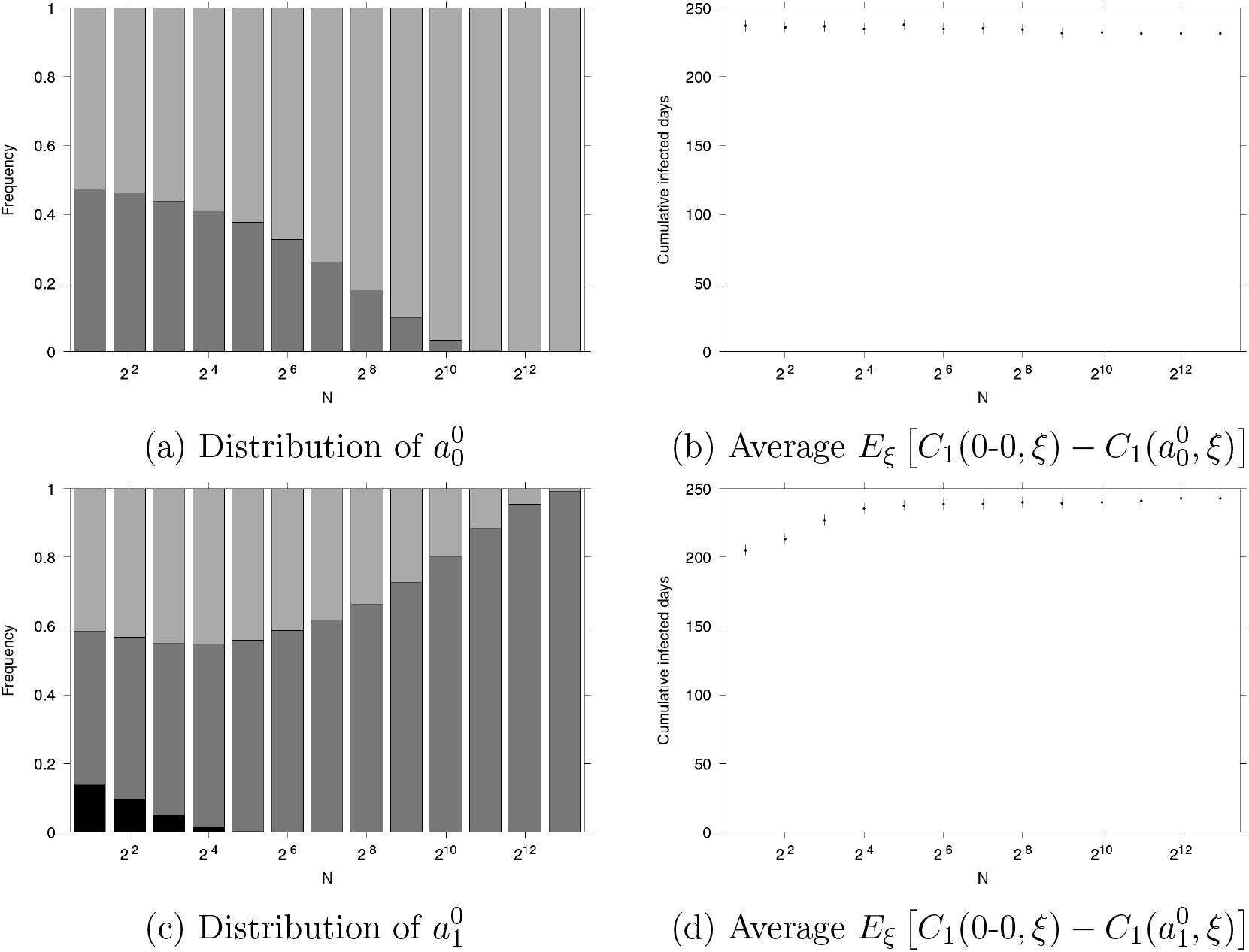
Decisions made under parameter uncertainty. Top panels: decisions based on *M*_0_. Bottom panels: decisions based on *M*_1_. Left panels: frequency of the policy picked, as a function of *N*. Black: no vaccination, dark grey: 0-50 policy, light grey: 50-0 policy. Right panels: average decision performance in averted symptomatic infected-days (vertical bars: 95 % CI). Policy distributions and average performances are estimated based on 65,536 decision making experiments.

Similarly, Figure 4c shows that decision 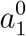 made based on stochastic model *M*_1_ under parameter uncertainty becomes less volatile as *N* increases and converges to 0-50, that is 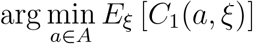 (Table 2). However, for small values of *N*, policy 0-0 (no vaccination) is sometimes picked due to stochastic effects (earlier extinction of the epidemic without vaccination than under policy 0-50 or 50-0 due to chance). Since policy 0-0 is actually suboptimal, the average performance of decisions made based on *M*_1_ is reduced for small values of *N* (Figure 4d).

### 3.3 Informed decision making

Let us now look into decision making when the true parameter values *ξ*\* are known. Recall that in this scenario, policy 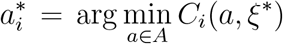 is picked based on model *M*_*i*_. Which policy performs better depends on parameter values *ξ*\* as shown in Figure 5. Policies 0-50 and 50-0 are optimal for about 50 % of parameter values each. Policy 0-0 (no vaccination) is optimal for no parameter values (Figure App-2 in Appendix).

**Figure 5:**
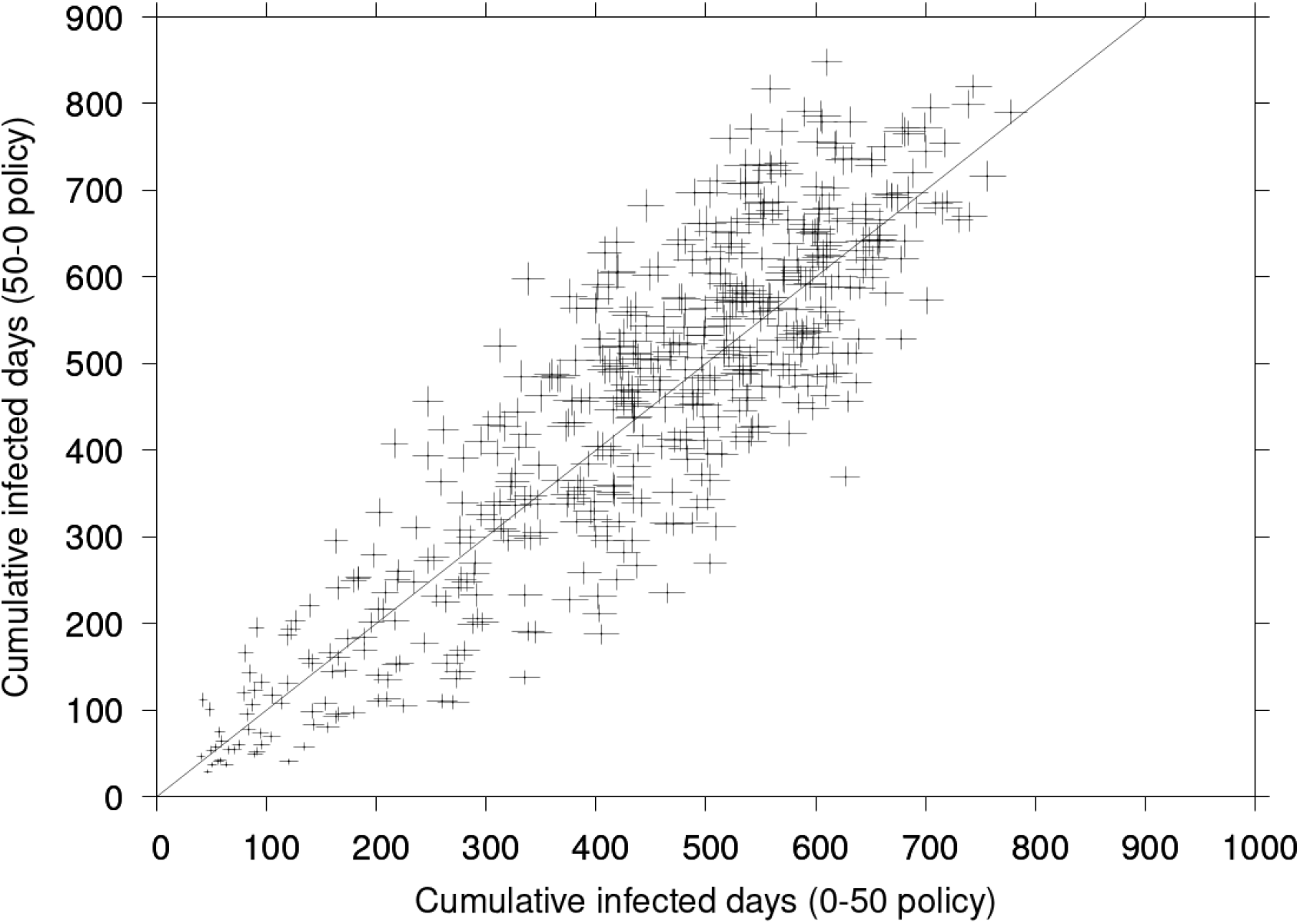
Cumulative symptomatic infected-days *C*_1_(0-50, *ξ*\*) (x-axis) and *C*_1_(50-0, *ξ*\*) (y-axis) for 512 data draws *ξ*\* from the base case parameter prior distributions. Crosses: 95% CI across 2,048 stochastic simulations of *M*_1_.

Figure 6 shows the influence of the number of model simulations *N* on decision making when the true parameter values *ξ*\* are known. When decisions are made based on deterministic model *M*_0_, decisions do not depend on the number of parameter runs (Figures 6a and 6b). Policies 0-50 and 50-0 are picked for about 50 % of parameter draws each (Figure 6a). When decisions are made based on stochastic model *M*_1_, we can observe the same stochastic effects as under uncertainty (Figure 4c): for small numbers *N* of model runs, suboptimal policy 0-0 is picked with non-zero probability (Figure 6c), which reduces the average performance of decision making (Figure 6d). As *N* increases, decisions converge towards 0-50 and 50-0 in almost equal proportion, consistently with Figure 5.

**Figure 6:**
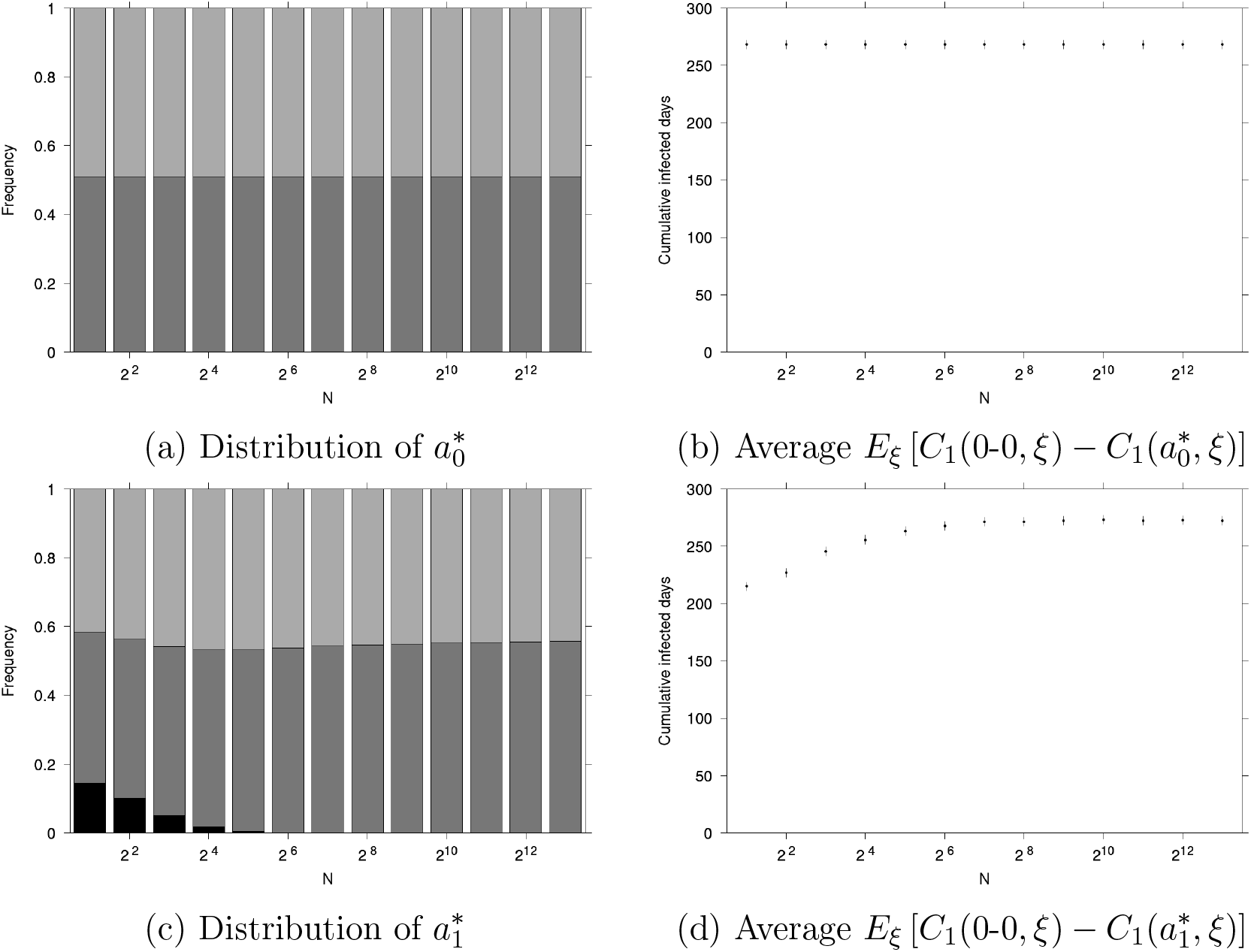
Decisions made knowing true parameter values. Top panels: decisions based on *M*_0_. Bottom panels: decisions based on *M*_1_. Left panels: frequency of the policy picked, as a function of *N*. Black: no vaccination, dark grey: 0-50 policy, light grey: 50-0 policy. Right panels: average decision performance in averted symptomatic infected-days (vertical bars: 95 % CI). Policy distributions and average performances are estimated based on 65,536 decision making experiments.

### 3.4 Cost of parameter uncertainty v. of using the wrong model

We showed that using a stochastic model where appropriate rather than its deterministic counterpart does influence the performance of decision making. We also showed that the number *N* of model runs can influence decision making as well, in particular when using a stochastic model. These effects hold under parameter uncertainty and when parameter values are known. Hence the question: how does the cost of using a deterministic model compares to that of deciding under uncertainty? And how does the number of model runs come into play in this respect?

Figure 7a shows the difference in average performance between decisions 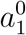 and 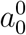 made under uncertainty based on stochastic model *M*_1_ and on deterministic model *M*_0_ respectively. It corresponds to the difference between Figures 4b and 4d. For small values of *N*, deterministic model *M*_0_ outperforms stochastic model *M*_1_ on average due to stochastic effects. As *N* increases, the difference converges to about 12 less cumulative symptomatic infected-days on average when decision are made with *M*_1_ rather than *M*_0_. In the context of decision making under uncertainty, *N* can be understood as a computational budget. Given this budget, it is possible i) to better sample the parameter prior distribution and use deterministic model *M*_0_ (one run per parameter draw), or ii) to use stochastic model *M*_1_ which is a more appropriate representation of reality but requires several runs for each parameter draw. This interpretation is of course limited since the computational resources actually needed for a single model run may vary greatly depending on the numerical methods or, in the case of the stochastic model, on the number of random events until simulation end (in our case, epidemic extinction). Still, we show with an example that an appropriate stochastic model can be outperformed by its deterministic version if the number of model runs is too small. We leave accurate estimations of required computational resources outside the scope of the present article since this is largely problem specific. We also do not consider the technical issue of balancing parameter draws and model runs per draw when making decisions under uncertainty using a stochastic model. Notice finally that in our example, the threshold value of *N* under which *M*_0_ outperforms *M*_1_ is rather small (about 16 or 32 runs) considering the simplicity of our transmission model and optimization problem. This is clearly problem dependent. In our robustness checks (Appendix F), we easily found instances in which stochastic effects still had an influence on the average performance of 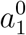 for *N* = 2^11^ = 2,048, and up to *N* = 2^13^ = 8,192 model runs. Besides, the maximum manageable number of model runs depends not only on the problem but also on available computing resources.

**Figure 7:**
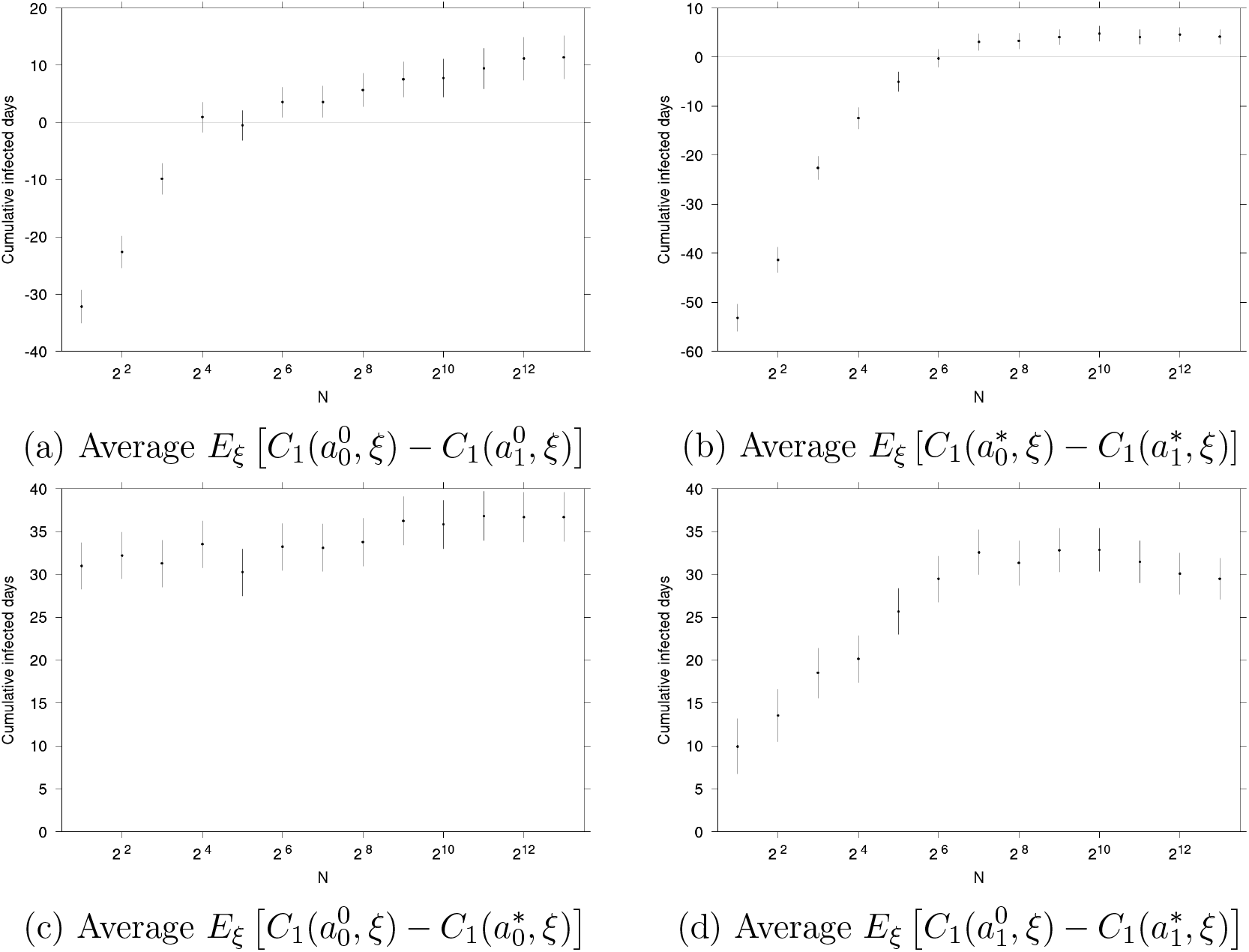
Differences in average decision making performance depending on the model used (*M*_0_ or *M*_1_) as functions of *N* and on the level of parameter information. Vertical bars: 95% CI.

Figure 7b shows the difference in average performance between decisions 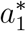 and 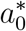 made knowing true parameter values based on stochastic model *M*_1_ and on deterministic model *M*_0_ respectively. It corresponds to the difference between Figures 6b and 6d. We can observe the same influence of stochastic effects for small values of *N* as in the scenario of decision making under uncertainty. Notice that given *N*, the benefit of using stochastic model *M*_1_ rather than deterministic model *M*_0_ is smaller in the informed decision making scenario than under the uninformed scenario (Figure 7a).

Figure 7c shows the additional benefit of resolving parameter uncertainties prior to making a decision, given that deterministic model *M*_0_ is used for decision making (comparison of 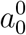 and 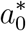). It corresponds to the difference between Figures 4b and 6b. In our example, policies 0-50 and 50-0 are picked with about 50 % chance for *N* = 2 both under parameter uncertainty and knowing parameter values (compare Figures 4a and 6a). Under parameter uncertainty, this is due to the poor sampling of the parameter prior distribution used to estimate *E*_*ξ*_ [*C*_0_(*a, ξ*)] for each policy *a*. For *N* = 2, decision making is essentially equivalent to picking a policy at random (among the non-dominated policies). In the informed case, 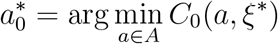 is policy 0-50 for about 50 % of the possible parameter values *ξ*\*, and policy 50-0 for the remaining parameter values. Policy 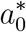 performs better than 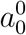 on average for *N* = 2 even though the distribution between policies 0-50 and 50-0 in each case are almost identical. The reason is that although deterministic model *M*_0_ is unable to capture some features of the epidemic, it still carries some information and allows to make better decisions knowing parameter values than those made under uncertainty.

The quantity 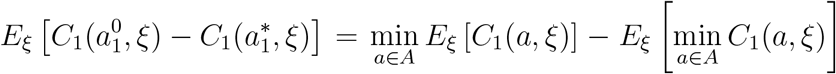 shown in Figure 7d is the expected value of perfect information (EVPI), a popular value-of-information metric which quantifies the additional benefit of entirely resolving uncertainties prior to making a decision [23]. It corresponds to the difference between Figures 4d and 6d. EVPI is positive or null by definition. It is subject to stochastic effects for small values of *N*.

We already showed that decisions made using the appropriate model, stochastic model *M*_1_, and in particular decisions 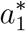 made based on *M*_1_ *and* knowing parameter values, can be outperformed due to stochastic effects for small values of *N*. Actually for *N* = 2, the best performance is achieved by policy 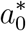 The performance of 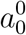 is decreased by using the “right” model (Figure 7a) but it can be improved by resolving uncertainties (Figure 7c). For large values of *N* however, the best performance is achieved by 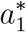. But still, starting from 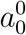, switching to model *M*_1_ and resolving uncertainties do not make the same contribution to improving performance: for *N* = 2^13^, switching to *M*_1_ allows to avert about 12 symptomatic infected-days (Figure 7a) against about 37 symptomatic infected-days averted by resolving uncertainties (Figure 7c).^2^

Results shown in Figure 7 are based on the baseline parameter prior distributions introduced in Section 2.2. As a robustness check we performed these analyses for randomly drawn prior distributions (the distribution parameters are drawn at random) and obtained qualitatively similar results, see Appendix F.

## 4 Conclusion

The respective properties of deterministic and stochastic infectious disease transmission models have been largely studied and are now introduced in most epidemiological modeling textbooks. By assuming discrete populations, stochastic models allow to account for effects that deterministic models cannot explain, typically (but not only) where epidemic extinction is involved. Yet the implications for decision making of the use of a deterministic or stochastic model have been somewhat overlooked in the literature. Some authors, it is true, compared the performance of vaccination policies picked based on a stochastic model with those picked based on its deterministic counterpart. However they did so mostly in order to provide a counterfactual to their stochastic optimization methods. They took the perspective of omniscient researchers investigating “from above” the relative performance of policies based on the stochastic and deterministic models in different areas of the parameter space. By contrast, we focused on the perspective of a decision maker. In particular we considered scenarios in which model parameters were uncertain, as is most often the case in real life.

Thus, we considered a total of four decision making scenarios: based on a stochastic model or its deterministic counterpart, and under uncertainty or knowing true parameter values. We looked into these scenarios using two minimal working examples of transmission models, a deterministic and a stochastic SIR models of an emerging disease, and a simple optimization problem consisting in picking a vaccination policies out of three alternative options (including a dominated no vaccination policy). The objective was to minimize the cumulative number of symptomatic infected-days over the course of the epidemic. In a validation step, we assessed the performance of the chosen policies by implementing them in the stochastic framework, which we assumed to be an accurate model of reality.

In our example, one of the non dominated policies was better at “nipping the epidemic in the bud” while the other was better at reducing the number of symptomatic infected-days in a large scale outbreak. Each policy was optimal for about 50 % of the possible parameter values. Under parameter uncertainty, a policy is picked that minimizes the expected number of cumulative symptomatic infected-days. We showed that in our case, different policies minimize the expected burden estimated by the deterministic model and the expected burden estimated by the stochastic model. This is because whether under parameter uncertainty or knowing parameter values, the deterministic model does not properly take the possibility of early epidemic extinction into account.

The performance of decision making largely depends on the quality of the expected performance estimates used to pick a policy. We then performed our analyses for different Monte Carlo sample sizes: this covers the size of parameter samples in the uniformed case, and the number of stochastic model runs per set of parameter values where the stochastic model is used. We showed that due to stochastic effects, the deterministic model can outperform the stochastic model for small sample sizes, both under parameter uncertainty or assuming informed decision making.

Our study comes with the usual caveats and provisions. Our aim was to provide an understanding of the interplay between the choice of a stochastic or deterministic model, parameter uncertainties, and Monte Carlo sample sizes from the perspective of decision making. Specific results presented here obviously depend on the transmission model structure or the parameter prior distributions. For instance our results suggest that resolving uncertainties brings more benefit than using the “right” (stochastic) model. We do not expect this to hold in all real life cases. We estimated the value of resolving uncertainties by assuming that true parameter values can actually be known. The value of information literature provides alternative approaches based on intermediate levels of information that may be more relevant in practice. As for our discussion of Monte Carlo sample sizes, it would ultimately need to consider specific numerical methods and available computational resources. We also left some technical issues such as finding the optimal balance between stochastic model runs and parameter sampling outside the scope of the study.

However, general results will remain true: the quality of decision making decreases with the sample size, stochastic models can be outperformed due to stochastic effects for small sample sizes, and this is true both under uncertainty and knowing parameter values. More importantly, the reasoning presented in the article will be relevant in any situation where stochastic effects are critical. The extra burden of sampling stochastic epidemic trajectories will have to be balanced against available resources and the cost of sampling the parameter space – this could be critical when solving a difficult optimization problem e.g. with heuristics requiring some fine tuning. Value-of-information metrics provide a useful benchmark when measuring the benefit of increasing the sample size or using a different model for decision making. In this, our study points to issues to be carefully considered when embarking on optimization of stochastic transmission models.

## Data Availability

All data produced in the present work are contained in the manuscript

## Appendix

### A Compartmental models

Let *S*_*i*_ and *I*_*i*_ denote the number of susceptible and infectious individuals who received *i* ∈ {0, 1, 2} doses of vaccine, and *R* the number of recovered individuals. Individuals get infected with contact rate *β* and they recover at rate *γ*. The basic reproduction number is *R*_0_ = *β/γ*. Individuals who received *i* ∈ {1, 2} vaccine doses are less susceptible by a factor *σ*_*i*_ and less infectious by a factor *ρ*_*i*_ compared to unvaccinated individuals. We assume that vaccination is instantaneous and that there is no waning of vaccine protection. In the deterministic model *M*_0_, the epidemic dynamics is described by Equations (1)–(5). The corresponding stochastic model *M*_1_ is summarized by events shown in Table App-1. Figure App-1 is a sketch of the transmission models.

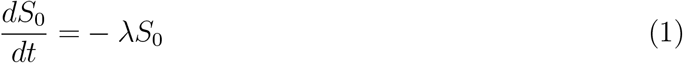

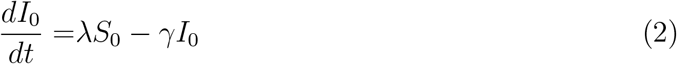

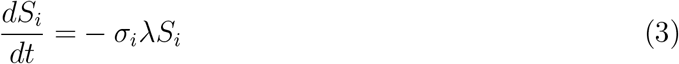

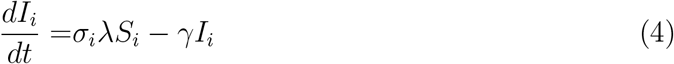

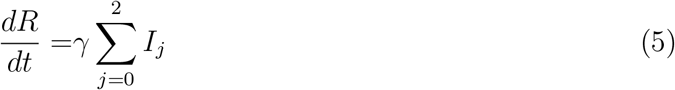

where

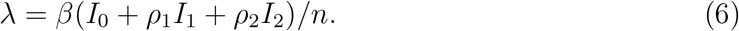

**Table App-1:** Events included in stochastic model *M*_1_.

| Event | Population changes | Rate |
| --- | --- | --- |
| Infection of unvaccinated | $S_0 \leftarrow S_0 - 1, I_0 \leftarrow I_0 + 1$ | $\lambda S_0$ |
| Infection of vaccinated | $S_i \leftarrow S_i - 1, I_i \leftarrow I_i + 1$ | $\sigma_i \lambda S_i$ |
| Recovery | $I_i \leftarrow I_i - 1, R_i \leftarrow R_i + 1$ | $\gamma I_i$ |

Let *p*_*i*_ the probability of symptomatic infection for infected individuals who received *i* doses of vaccine. The incidence rates of symptomatic and asymptomatic cases among unvaccinated individuals are 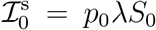 and 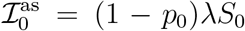 respectively. Among individuals who received *i* ∈ {1, 2} doses of vaccine, incidence rates are 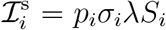 and 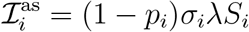.

**Figure App-1:**
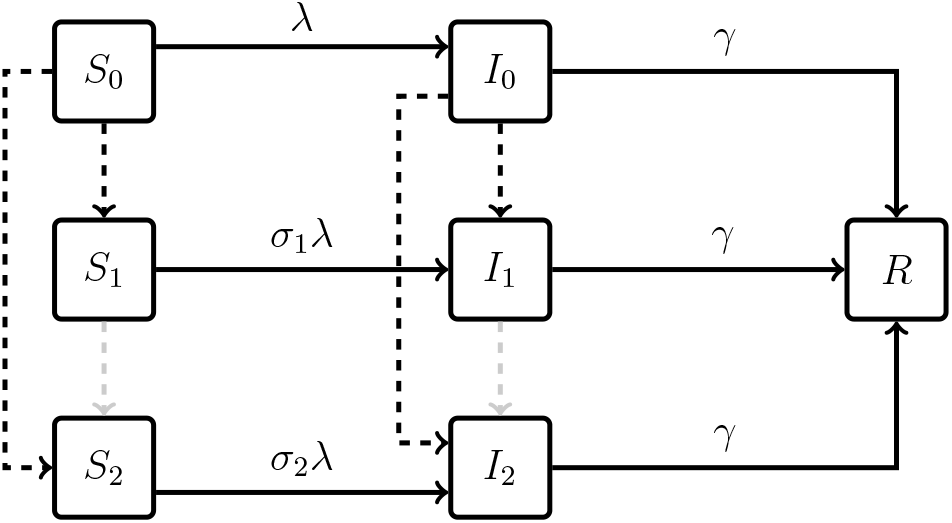
Sketch of the transmission models. Dashed: possible vaccination. Grey dashed: vaccination not considered in the present article.

### B Parameter prior distributions

For convenience, we use truncated normal distributions as parameter prior distributions. Table App-2 shows the mean and standard deviation of the underlying normal distributions, and the supports of the prior distributions. Once a set of parameter is drawn, we check that *p*_1_ > *p*_2_, *ρ*_1_ > *ρ*_2_ and *σ*_1_ > *σ*_2_. Hence, the reader should be aware that parameter values are not independent.

**Table App-2:** Base case parameter prior distributions.

| Parameter | Mean | Std. dev. | Support |
| --- | --- | --- | --- |
| $R_0$ | 4.0 | 4.0 | $[1.2, \infty[$ |
| $\gamma$ | 0.1 | 0.0 | — |
| $\sigma_1$ | 0.8 | 0.3 | $[0.02, 1.0]$ |
| $\rho_1$ | 0.8 | 0.3 | $[0.02, 1.0]$ |
| $p_1/p_0$ | 0.8 | 0.3 | $[0.02, 1.0]$ |
| $\sigma_2$ | 0.5 | 0.3 | $[0.02, 1.0]$ |
| $\rho_2$ | 0.5 | 0.3 | $[0.02, 1.0]$ |
| $p_2/p_0$ | 0.5 | 0.3 | $[0.02, 1.0]$ |

### C Stochastic effects: additional table

**Table App-3:** Estimate differences in expected outcome between the three available policy options based on *M*_0_. Averages over 2^20^ parameter draws (one run of *M*_0_ per parameter draw).

| **Metric** | **Estimate** |
| --- | --- |
| $E_{\xi} [C_0(0-0, \xi) - C_0(0-50, \xi)]$ | 215.064 (95% CI: 214.947 – 215.18 ) |
| $E_{\xi} [C_0(0-0, \xi) - C_0(50-0, \xi)]$ | 215.359 (95% CI: 215.122 – 215.597 ) |
| $E_{\xi} [C_0(0-50, \xi) - C_0(50-0, \xi)]$ | 0.295686 (95% CI: 0.100199 – 0.491173) |

### D Bootstrapping procedures

We bootstrap our simulations (512 parameter draws and 2,048 stochastic simulations per available policy for each data draw) in order to obtain 65,536 total simulations of decision making.

For decision making based on deterministic model *M*_0_ under parameter uncertainty, the bootstrapping procedure is as follow:

- 65,536 times do:
  – draw parameter values *ξ*\*,
  – *N* times do:
    * draw parameter values *ξ*,
    * for each policy *a ∈ A*, compute *C*_0_(*a, ξ*),
  – for each policy *a ∈ A*, estimate *E*_*ξ*_ [*C*_0_(*a, ξ*)] based on the *N* parameter draws *ξ*,
  – pick 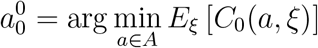 based on these estimates,
  – validation: compute 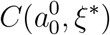 (one run of stochastic model *M*_1_),
- estimate 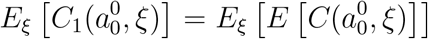 based on the 65,536 simulations of decision making.

For decision making based on stochastic model *M*_1_ under parameter uncertainty, the bootstrapping procedure is as follow:

- 65,536 times do:
  – draw parameter values *ξ*\*,
  – *N* times do:
    * draw parameter values *ξ*,
    * for each policy *a* ∈ *A*, compute *C*(*a, ξ*) (one run of *M*_1_ for each policy),
  – for each policy *a* ∈ *A*, estimate *E*_*ξ*_ [*C*_1_(*a, ξ*)] = *E*_*ξ*_ [*E* [*C*(*a, ξ*)]] based on the *N* runs of *M*_1_ and parameter draws *ξ*,
  – pick 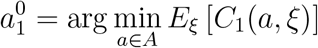 based on these estimates,
  – validation: compute 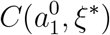 (one run of stochastic model *M*_1_),
- estimate 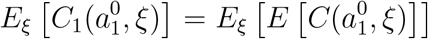 based on the 65,536 simulations of decision making.

For decision making based on deterministic model *M*_0_ knowing parameter values, the bootstrapping procedure is as follow:

- 65,536 times do:
  – draw parameter values *ξ* *,
  – for each policy *a* ∈ *A*, compute *C*_0_(*a, ξ*\*),
  – pick 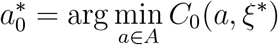,
  – validation: compute 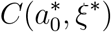 (one run of stochastic model *M*_1_),
- estimate 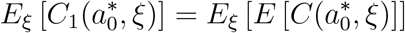 based on the 65,536 simulations of decision making.

For decision making based on stochastic model *M*_1_ knowing parameter values, the bootstrapping procedure is as follow:

- 65,536 times do:
  – draw parameter values *ξ* *,
  – *N* times do:
    * for each policy *a* ∈ *A*, compute *C*(*a, ξ* *) (one run of *M*_1_ for each policy),
  – for each policy *a* ∈ *A*, estimate *C*_1_(*a, ξ* *) = *E* [*C*(*a, ξ* *)] based on the *N* runs of *M*_1_,
  – pick 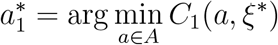 based on these estimates,
  – validation: compute 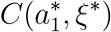(one run of stochastic model *M*_1_),
- estimate 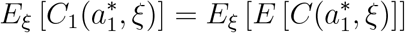 based on the 65,536 simulations of decision making.

### E Informed decision making: additional figures

**Figure App-2:**
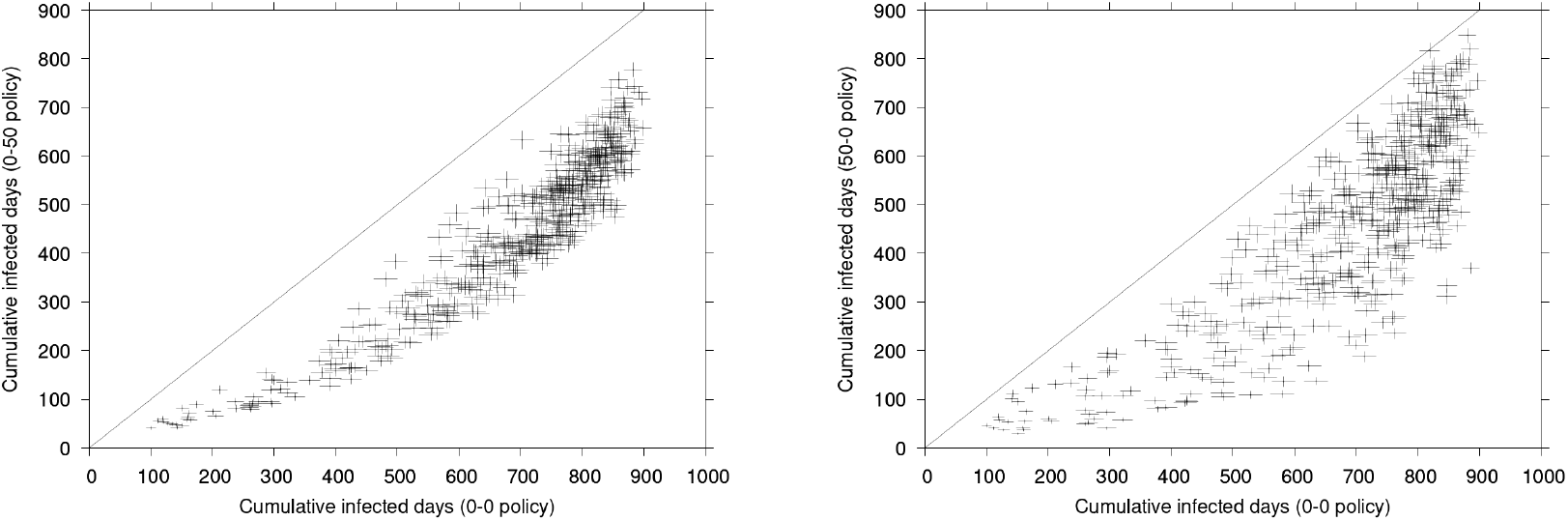
Cumulative symptomatic infected-days *C*_1_(0-0, *ξ*\*) (x-axis) and *C*_1_(0-50, *ξ*\*) (y-axis, left panel) or *C*_1_(50-0, *ξ*\*) (y-axis, right panel) for 512 data draws *ξ*\* from the base case parameter prior distributions. Cross heights and widths represent 95% CI across 2,048 stochastic simulations of *M*_1_.

### F Robustness checks

In Figure App-3, we show results similar to those presented in Figure 7 for different priors drawn by chance as displayed in Table App-4.

**Table App-4:** Distributions of the parameters of parameter prior distributions. *U* (*a, b*): uniform distribution with support [*a, b*]. 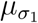: mean parameter (mean of the underlying normal distribution) of the prior distribution of *σ*_1_.

| Parameter | Mean | Std. dev. |
| --- | --- | --- |
| $R_0$ | $U(2, 10)$ | $U(1, 5)$ |
| $\gamma$ | $U(0.1, 0.5)$ | $U(0, 0.2)$ |
| $\sigma_1 = \rho_1 = p_1/p_0$ | $U(0.5, 0.9)$ | $U(0.1, 0.4)$ |
| $\sigma_2 = \rho_2 = p_2/p_0$ | $U(0.1, \mu_{\sigma_1})$ | $U(0.1, 0.4)$ |

**Figure App-3:**
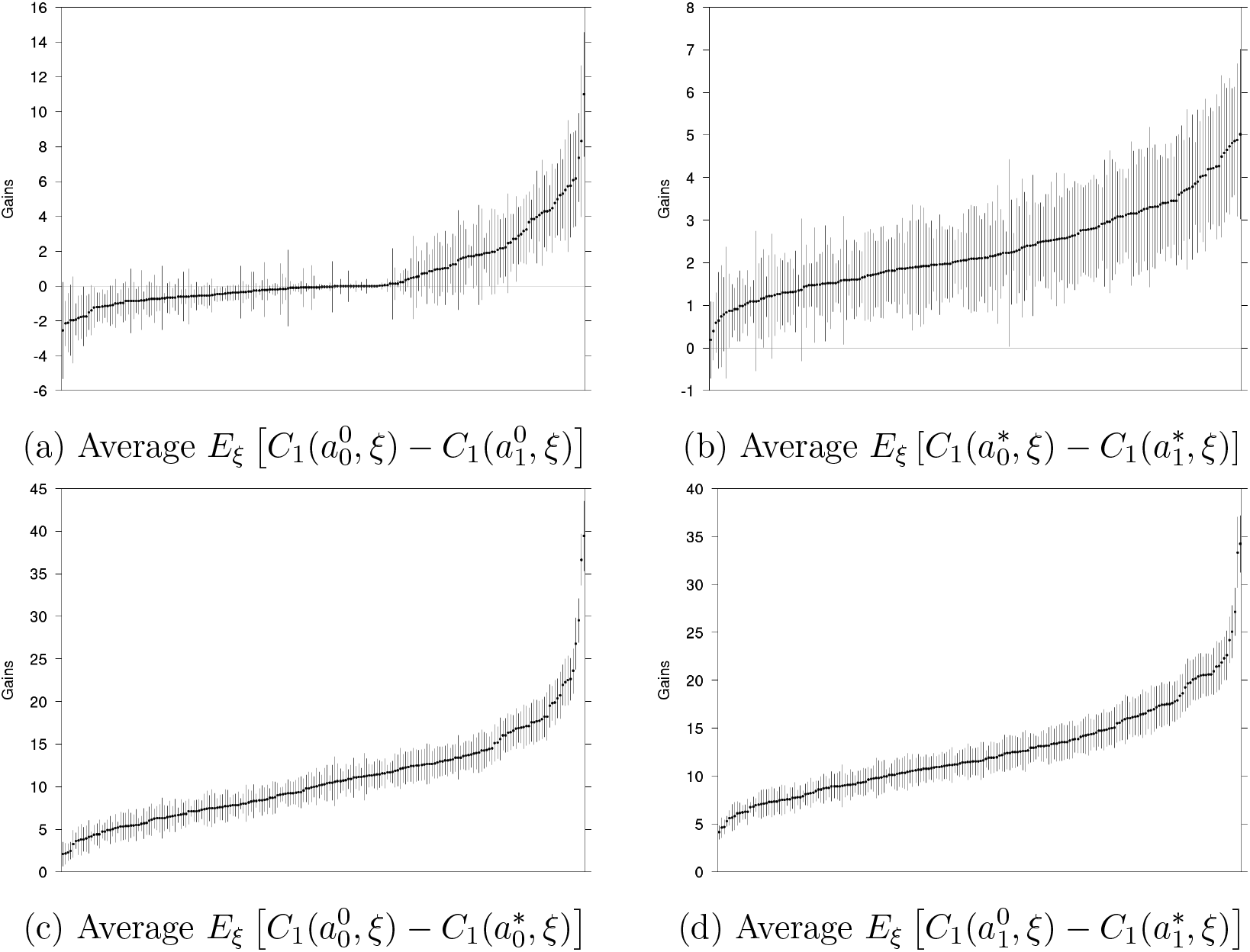
Differences in cumulative symptomatic infected-days between different cases for prior distributions drawn by chance as described in Table App-4. Results are ordered by increasing value. Vertical bars: 95% CI. *N* = 2, 048.

Remark: the fact that many values of the average performance difference 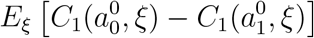 between *M*_0_ and *M*_1_ under uncertainty (Figure App-4a) are negative for *N* = 2, 048 = 2^11^ seems to be due to *N* being too small. In figure App-4, we display the same as Figure 7 for the set of values with the second lowest average value in Figure App-4a (the smallest is not statistically significant). Figure App-5 shows the corresponding decision distributions. We see from Figure App-5c that stochastic effects still play a role for *N* = 2^1^3 = 8,192 as the distribution of 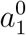 has not fully converged.

**Figure App-4:**
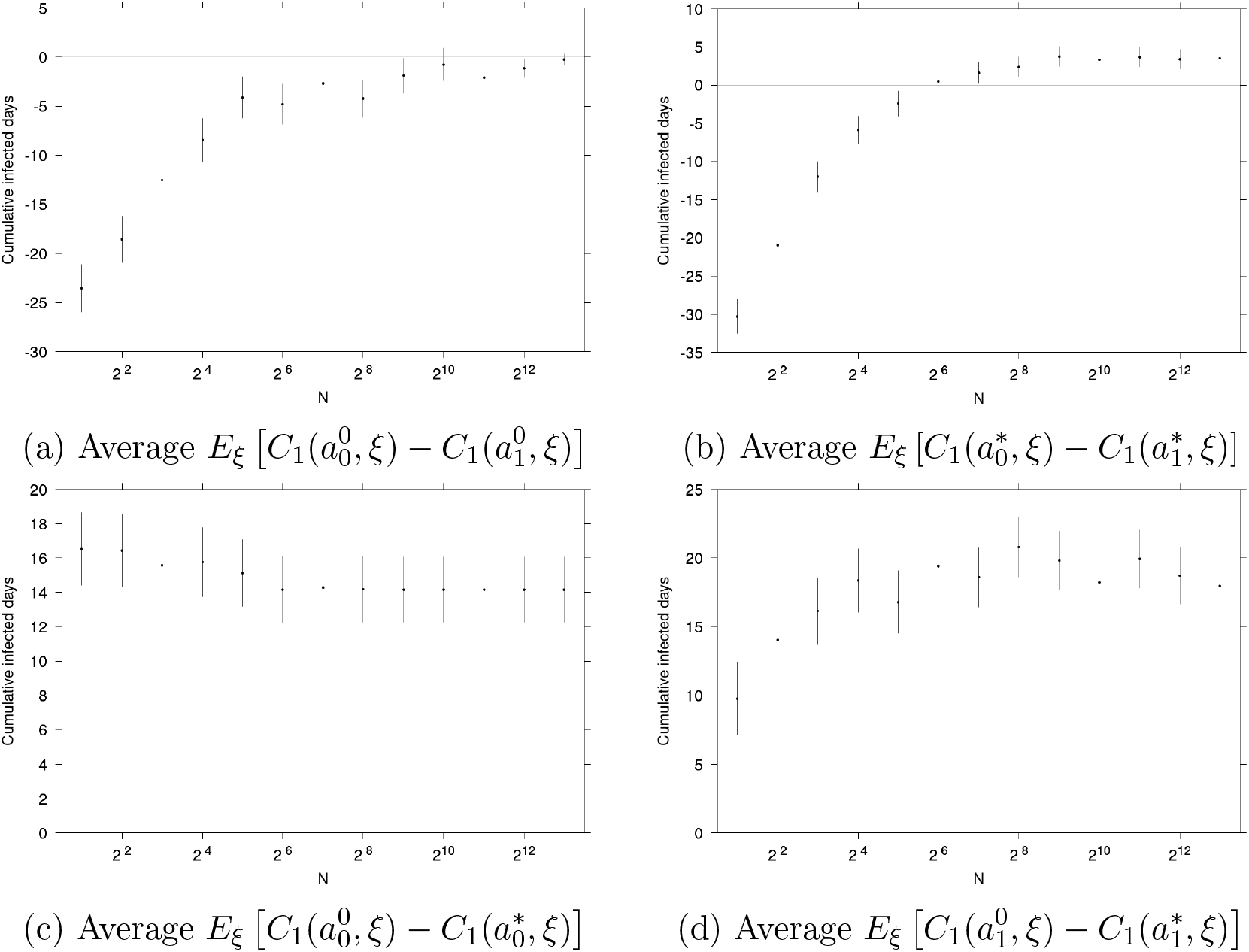
Difference between numbers of cumulative infected days avoided in the different cases and as functions of *N*. Vertical bars: 95% CI.

**Figure App-5:**
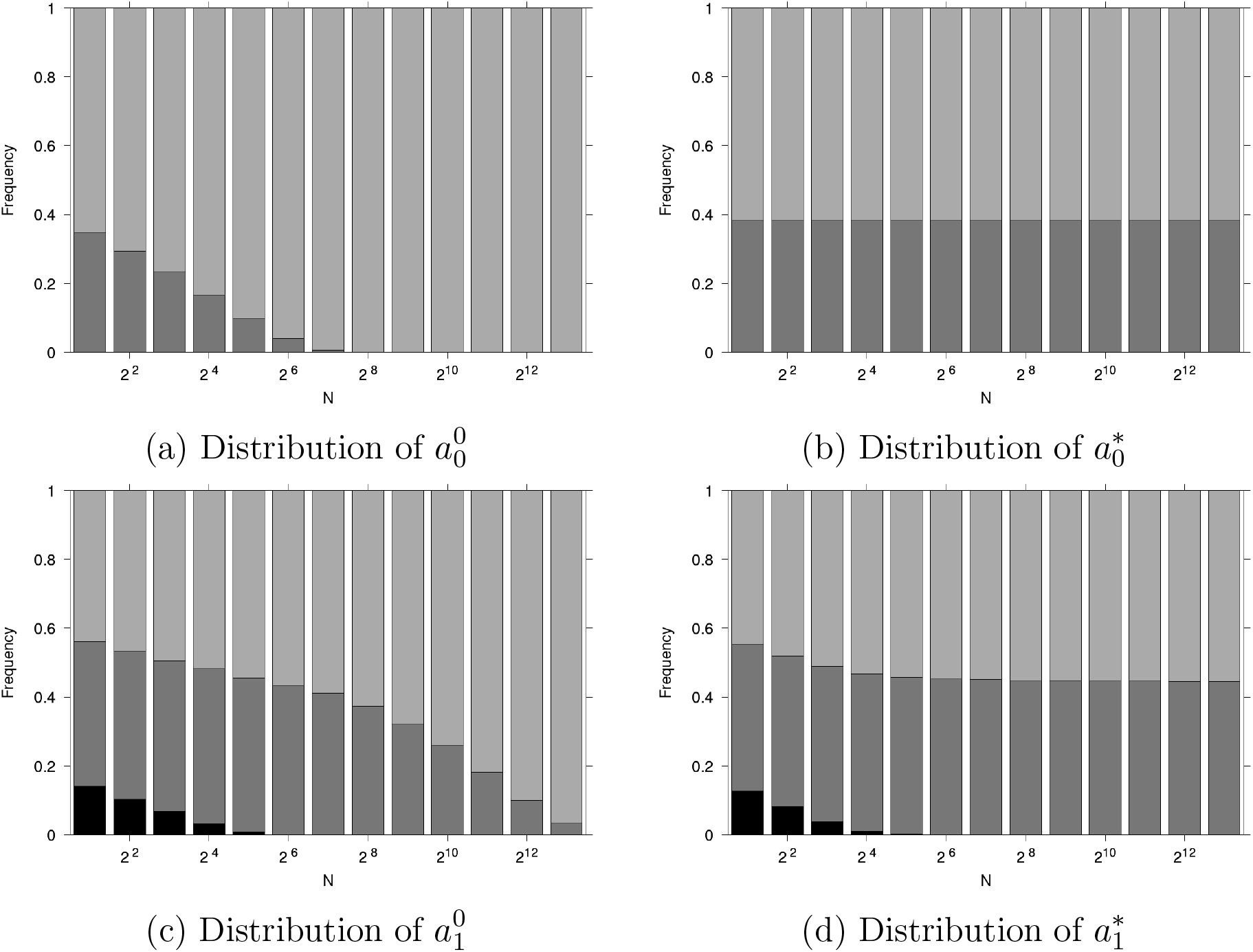
Frequency of the policy implemented as a function of *N*. Black: no vaccination, dark grey: 0-50 policy, light grey: 50-0 policy.

## Footnotes

1 Notice that confidence intervals are displayed for the sake of completeness. However, they do not inform on statistical significance of the policy *differences* per parameter values. Relevant confidence intervals are given in the remainder of the text. We provide estimates of expected outcome differences based on *M*_0_ in Table App-3 in Appendix, although this will not bear on our resoning.

2 For this value of *N*, all differences have converged to their large sample size values.

